# Factors affecting the mental health of pregnant women using UK maternity services during the COVID-19 pandemic: A qualitative interview study

**DOI:** 10.1101/2021.10.20.21265279

**Authors:** A R McKinlay, D Fancourt, A Burton

## Abstract

**Background:** People using maternity services in the United Kingdom (UK) have faced significant changes brought on by the COVID-19 pandemic and social distancing regulations. We focused on the experiences of pregnant women using UK maternity services during the pandemic and the impact of social distancing rules on their mental health and wellbeing.

**Methods:** We conducted 23 qualitative semi-structured interviews from June 2020 to August 2021, with women from across the UK who experienced a pregnancy during the pandemic. Nineteen women in the study carried their pregnancy to term and four women experienced a miscarriage during the pandemic. Interviews took place remotely over video or telephone call, discussing topics such as mental health during pregnancy and use of UK maternity services. We used reflexive thematic analysis to analyse interview transcripts.

**Results:** We generated six higher order themes: (1) Some pregnancy discomforts alleviated by social distancing measures, (2) The importance of relationships that support coping and adjustment, (3) Missed pregnancy and parenthood experiences, (4) The mental health consequences of birth partner and visitor restrictions, (5) Maternity services under pressure, and (6) Lack of connection with staff. Many participants felt a sense of loss over a pregnancy experience that differed so remarkably to what they had expected because of the pandemic. Supportive relationships were important to help cope with pregnancy and pandemic-related changes; but feelings of isolation were compounded for some participants because opportunities to build social connections through face-to-face parent groups were unavailable. Participants also described feeling alone due to restrictions on partners being present when accessing UK maternity services.

**Conclusions:** Our findings highlight some of the changes that may have affected pregnant women’s mental health during the COVID-19 pandemic. Reduced social support and being unable to have a partner or support person present during maternity service use were the greatest concerns reported by women in this study, as this absence removed a protective buffer in times of uncertainty and distress. This suggests that the availability of a birth partner or support person must be prioritised wherever possible to protect the mental health of women experiencing pregnancy and miscarriage in times of pandemics.

## Background

Pregnancy is a significant life stage tied to events that can affect mental health and wellbeing.(1) Relationship difficulties,(2) childcare responsibilities,(3) financial hardship,(4) stressful life events,(5) and natural disasters, (6) can also contribute towards maternal mental ill health. Many women not only experience profound changes in their identity, self-concept and sense of meaning during parenthood but can also experience complex emotions such as shame and guilt, (7) which makes the availability of support during this life stage so important. Up to 63% of new mothers are estimated to experience some ongoing symptoms of depression after birth, (8) with 10-15% experiencing symptoms sufficient for a diagnosis of postnatal depression. (9) The health and wellbeing of mothers during pregnancy is a critical public health issue as this period can pave the way for long-term health outcomes for parents and their children.(10)

When the World Health Organization declared coronavirus disease 2019 (COVID-19) a pandemic in March 2020,(11) the UK government advised pregnant women to take extra caution to protect themselves from infection.(12) The UK National Health Service also classified pregnant women as “clinically vulnerable” to the effects of COVID-19, (13). In response to the pandemic, the biggest changes to UK maternity care included face-to-face service cancellations, two-metre social distancing during appointments, restrictions of partners from attending outpatient appointments, and limiting the number of visitors during intrapartum care.(14) Although the measures were introduced to reduce the spread of the virus and protect those identified as clinically vulnerable, at present, little evidence has been published on the social and mental health impact of these changes during the various stages of the COVID-19 pandemic.

Experts warned early on that steps taken to reduce the risk of virus transmission from mothers with confirmed or suspected COVID-19 to their newborns may have indirect health consequences (15), such as difficulties with lactation and reduced mother-child bonding. (14,16) Unfortunately, such predictions have already been suggested to have been valid. Emerging research has found increases in depression, anxiety and loneliness amongst women in the perinatal period during the pandemic (17) and an experience of miscarriage may place people at potentially higher risk of these symptoms. (18) Health service changes due to the pandemic (such as missed appointments or cancelled services) have been found to be significantly associated with trauma symptoms, depression, anxiety and loneliness.(19) Karavadra et al. (2020) found many women were also concerned about remote antenatal appointments, partner visiting restrictions, and rapidly changing rules that affect health service provision.(20) In a qualitative interview study that explored barriers to healthcare seeking among women during the first UK lockdown in March 2020, authors found that women delayed seeking care due to fears of COVID exposure, negative media reports, and influence of social contacts. (21) Basu et al. (2021) also found that people experiencing pregnancy had concerns with changes in delivery plans and about the risk of their newborn catching COVID-19.(19)

While quantitative data suggests there have been increases in psychological distress for people experiencing pregnancy during the pandemic, (19) limited qualitative evidence has been published on *why* women experienced a deterioration in mental health during this time. One qualitative study identified increased feelings of isolation and difficulties accessing breast-feeding and parenting support. (3) However, this study focused specifically on the postpartum period and experiences of women who had given birth, either before or during the very early stages the pandemic. Furthermore, it did not include the views of women experiencing a miscarriage during this time. In this research, we sought to learn more about changes across the COVID-19 pandemic that may have contributed to a decline in mental health and wellbeing amongst pregnant women.

## Methods

### Study Design

We used in-depth qualitative interview methods to elicit experiences and perspectives of 23 women using UK maternity services during the COVID-19 pandemic. Our aims were to explore women’s experiences of maternity service use during the COVID-19 pandemic and describe how these experiences affected mental health and wellbeing. We did this by exploring in what ways the pandemic has affected pregnant women’s mental health, wellbeing and subjective experiences through one-off interviews conducted remotely over video or mobile phone call. We obtained ethical approval prior to undertaking the study from the University College London Ethics Committee (Project ID: 14895/005).

### Recruitment

We recruited a convenience sample of participants by circulating advertisements through social media (i.e., Twitter), a study newsletter (reaching around 3,000 people), and personal contacts. Interested people contacted the research team to register for the study. AM or AB responded with further information and a screening questionnaire. Everyone who registered their interest had the opportunity to ask questions before joining the study. Eligibility criteria included being 18 years or older, having experienced a pregnancy and accessed UK maternity services during the pandemic and being able to speak English sufficiently to read and understand the study information and informed consent forms.

In the context of an ongoing pandemic and choice of study population where demographic factors can have important implications for participant experiences,(22) we opted to use some purposive sampling strategies during study recruitment, whereby we screened participants in attempt to ensure diversity within the group. The factors of interest during study recruitment known to affect pregnancy outcomes included maternal age, education level,(23) and ethnicity.(24,25) In considering the potential mental health impact of miscarriage (26) we also invited participants to take part based on experiencing a pregnancy and use of UK maternity services during the pandemic, rather than giving birth and carrying to full term alone.

### Procedure

Interviewers (authors AM, AB) were female, PhD-level, qualitative health researchers with training in mental health. Participants completed their interviews from June 2020 until August 2021. We offered participants a remote interview via telephone or online video call. Participants provided informed written consent before taking part and completed a demographics form. Interviews followed a topic guide designed to illicit responses on pregnancy experiences during the pandemic and the impact on mental health, wellbeing, access to support and social lives. See Table 1 for examples of questions asked.

**Table 1.**
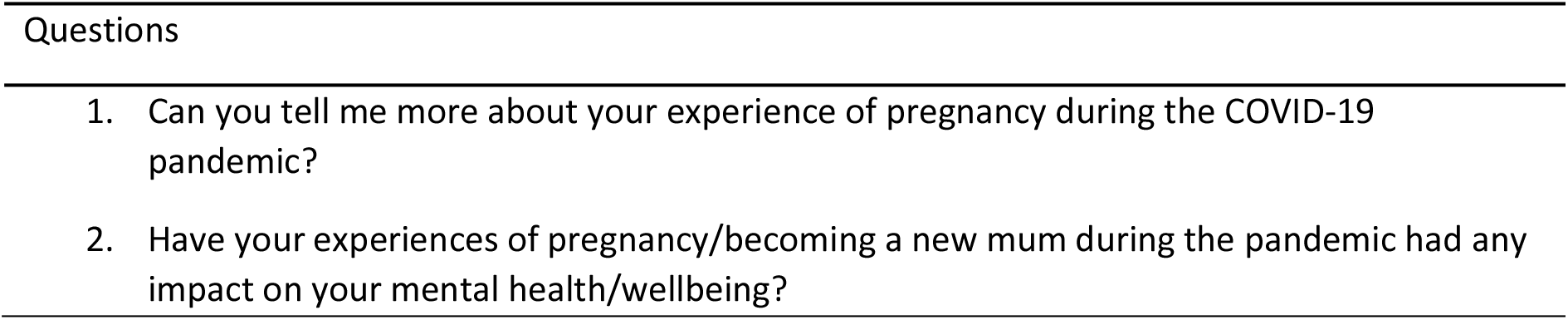
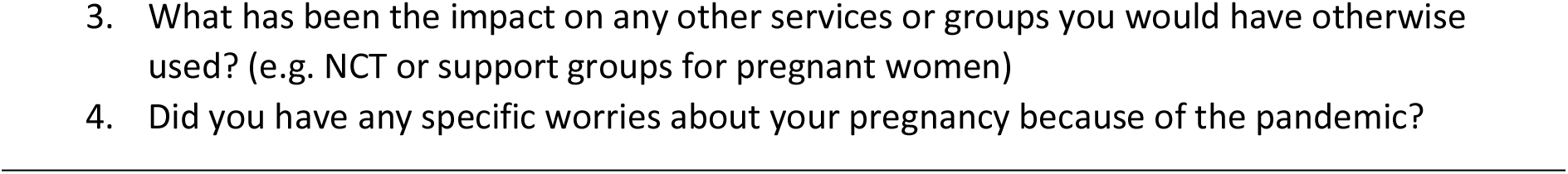
Topic Guide Examples

### Data Analysis

Audio files from participant interviews were transcribed verbatim by a third-party transcription service. AB and AM checked transcripts for accuracy and anonymity before importing into Nvivo version 12.(27) AM led on data analysis with reference to reflexive thematic analysis (RTA) techniques (28,29) informed by critical realist ontology. (30) First, AM and AB independently coded three transcripts and met to discuss issues of importance identified in the transcripts. This step was carried out, not to generate a reliable coding framework, which RTA does not prescribe, (28) but to develop a more nuanced and contextualised approach for interpreting and coding the remainder of files. The coding framework was grounded in the data rather than being based on a pre-existing theory or structure. The lead author (AM) read all remaining transcripts and then coded these with a focus on concepts relevant to the research question, rather than line-by-line coding of all interview data. (29) AM generated themes and subthemes with input from co-authors (AB and DF). The preliminary findings were also presented to the CSS qualitative research team, a group of researchers who have used RTA to analyse previous work on the mental health impact of the pandemic among specific groups.(31–33)

## Results

### Participant Characteristics

We recruited 23 women who experienced a pregnancy and used UK maternity services during the COVID-19 pandemic (Table 2). Most participants described themselves as married, female and living with their partner. Fifteen women had one child, four women had two children, three women had no children following an experience of miscarriage (four women in total had experienced miscarriage during the pandemic, one of whom later gave birth to her first child), and one woman had three children. Sixty-one percent of participants identified as White British. Eight participants had a diagnosed pre-existing mental health condition, including premenstrual dysphoric disorder, depression, and anxiety. Two women were experiencing post-natal depression and anxiety at the time of their interview. Three women had a pre-existing physical health condition, including a mobility condition, skin condition, and polycystic ovarian syndrome.

**Table 2.**
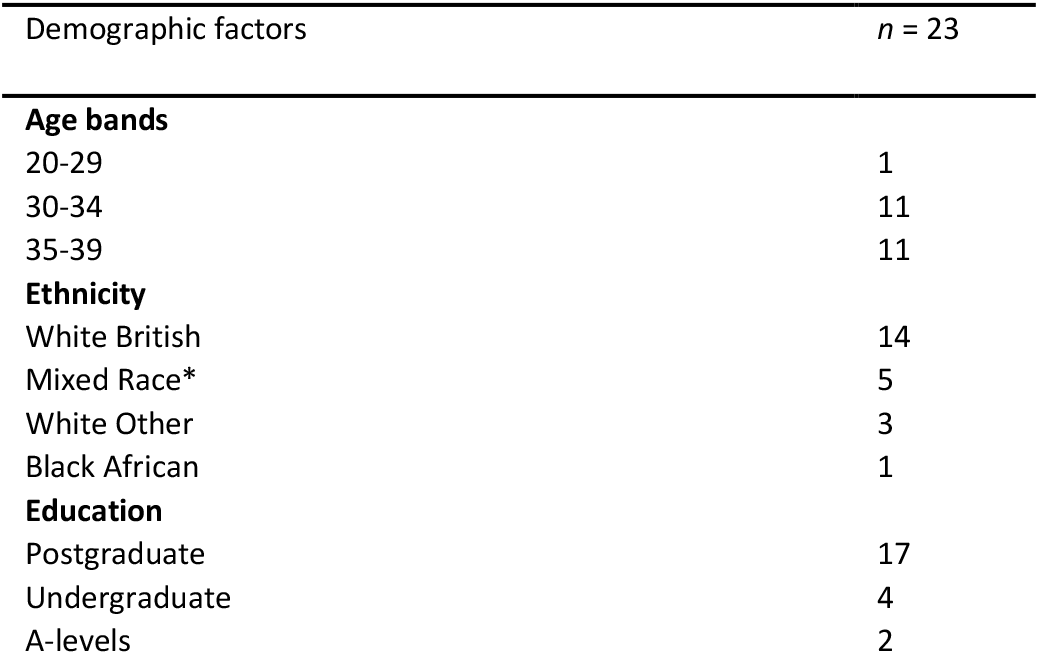
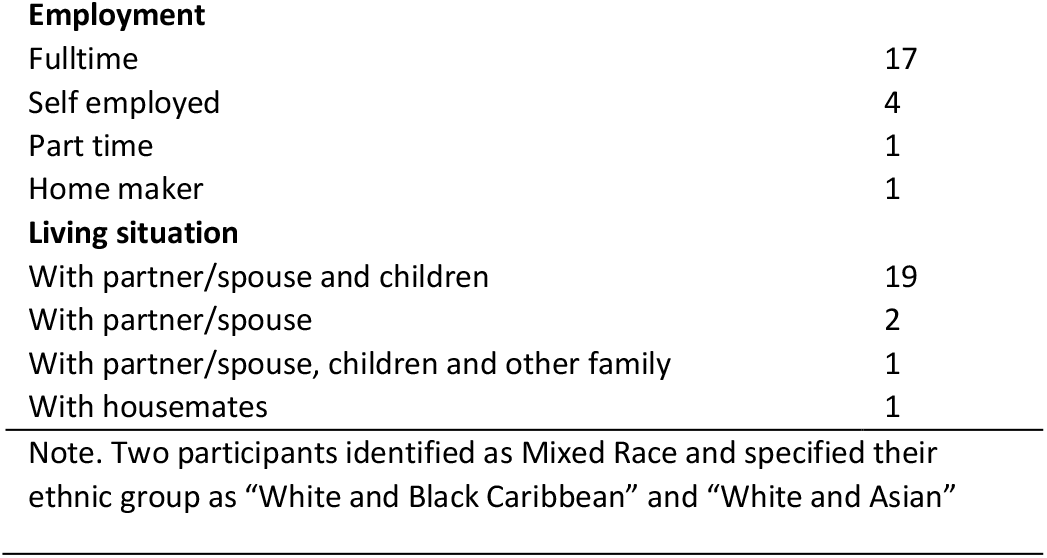
Participant characteristics

| Demographic factors | *n* = 23 |
| --- | --- |
| **Age bands** |  |
| 20-29 | 1 |
| 30-34 | 11 |
| 35-39 | 11 |
| **Ethnicity** |  |
| White British | 14 |
| Mixed Race* | 5 |
| White Other | 3 |
| Black African | 1 |
| **Education** |  |
| Postgraduate | 17 |
| Undergraduate | 4 |
| A-levels | 2 |
| **Employment** |  |
| Fulltime | 17 |
| Self employed | 4 |
| Part time | 1 |
| Home maker | 1 |
| **Living situation** |  |
| With partner/spouse and children | 19 |
| With partner/spouse | 2 |
| With partner/spouse, children and other family | 1 |
| With housemates | 1 |
| Note. Two participants identified as Mixed Race and specified their ethnic group as “White and Black Caribbean” and “White and Asian” |  |

### Thematic Analysis

We generated six themes during the analysis about the mental health impact of pandemic-related experiences of pregnant women in the UK (Table 3). Some of the emotions reported by women corresponded to their experiences during pregnancy or postpartum specifically, and others spanned across all stages of pregnancy and parenthood. No experiences were universal, and we have attempted to capture these nuances within the following discussion of themes and subthemes.

**Table 3.**
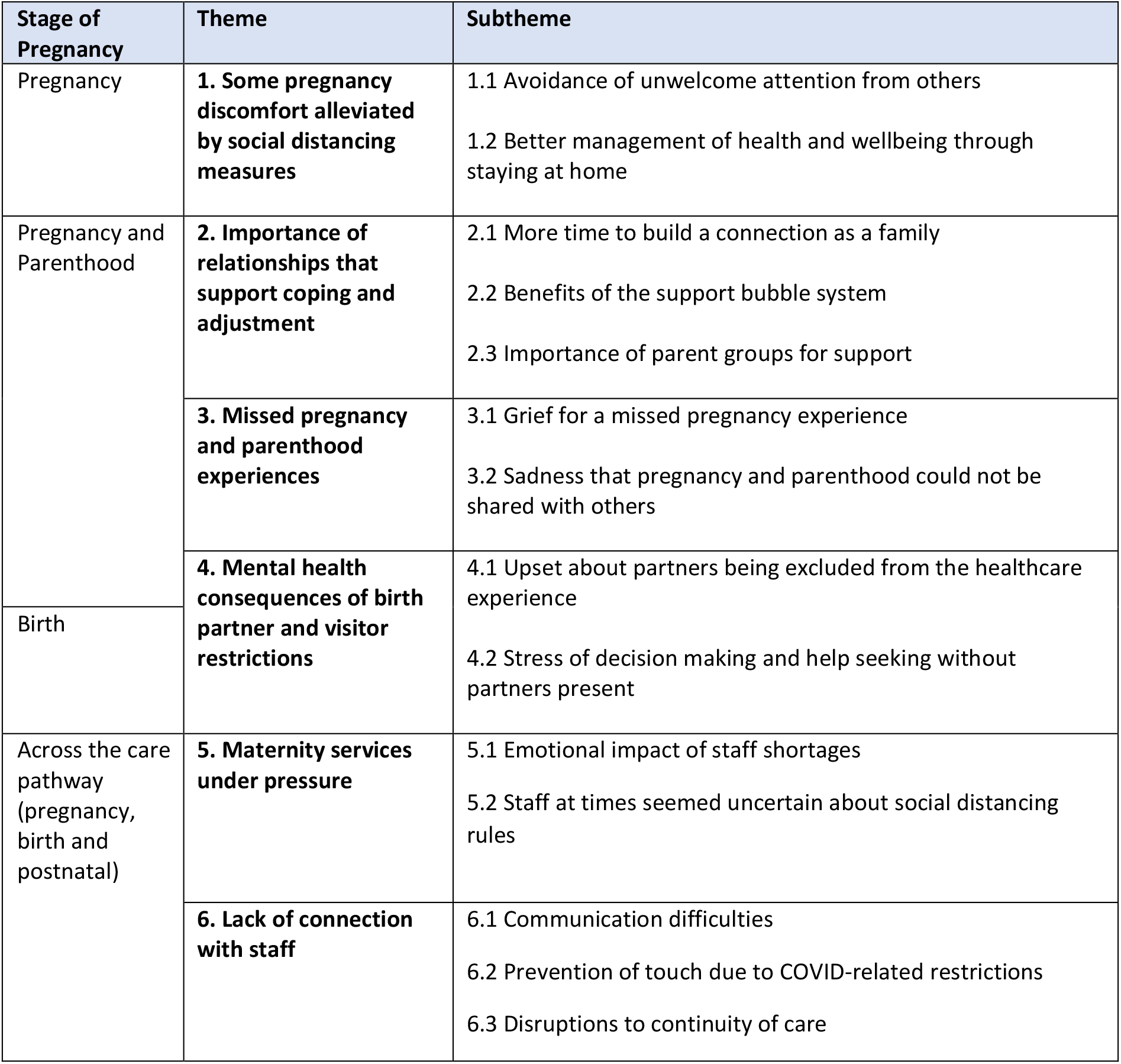
Summary of themes and subthemes described by participants.

## 1 Some pregnancy discomfort alleviated by social distancing measures

Most participants described some benefits during their pregnancy and parenthood that were associated with social distancing restrictions arising from the COVID-19 pandemic.

### 1.1 Avoidance of unwelcome attention from others

Social isolation measures meant some participants were able to experience changes in their body during pregnancy without the unwanted gaze, touch, or commentary of other people. One participant described feeling uncomfortable during a previous pregnancy when people touched her stomach. The requirement to physically distance from other people meant no one touched her without her expressed permission.

> *“I did not see anybody; nobody touched my bump. I remember being driven insane in my first pregnancy. Because my manager, who is very lovely… Used to touch my bump, every time she saw me, without asking*.*” P7, aged 35-40, 2*^*nd*^ *baby*

Social distancing regulations also meant that participants could avoid uncomfortable social encounters with others: “*It’s been nice not to have unsolicited visits, so that’s one pro of it*.” At other times, participants were able to avoid unwelcome discussions about the early stages of their pregnancy, and rather, initiate these conversations at their own pace when they felt comfortable.

> *“…especially when you’re not telling people that you’re pregnant, to be able to just do that and never have to tell someone I wasn’t drinking because I didn’t see anyone, so that was great*.*” P5, aged 30-34, no children, with experience of miscarriage during pandemic*

Some participants described this as helpful, particularly by women with pre-existing anxiety, high-risk pregnancy, and history of miscarriage.

> *“I think in a way not being able to see people has helped me a little bit. I think there have been times when I’ve been really, really low in the last year and a half and it’s been nice not to have to go and pretend that I’m all right… it’s been really nice not seeing anyone and just keeping it low key*.*” P22, aged 35-40, 1*^*st*^ *baby + miscarriage during pandemic*

### 1.2 Better management of health and wellbeing through staying at home

With the exception of participants working in frontline professions who were unable to continue face-to-face work, the introduction of lockdown measures meant many participants worked from home during their pregnancy. This change in routine meant that participants said they felt better equipped to manage the tiredness and nausea they experienced during early pregnancy.

> *“I would throw up and then I would just carry on, or I would be very tired, and I would lie down and then I would go back to work… And I was thinking, if it had been normal times, I would have had to take loads of time sick, off work, because I wouldn’t have been able to face getting on a train. The time I would have been on a train, is the time I threw up every day…” p12, aged 35-40, 1*^*st*^ *baby*

Some participants reported that being at home during lockdown resulted in noticeable health benefits. For example, they said they benefitted from not becoming sick with seasonal illness, as they might normally have done throughout the year.

> *“There was some benefits to social distancing, and like I said, we all benefited in terms of not getting sick*.*” P7, aged 35-40, 2*^*nd*^ *baby*

Health improved for some during their pregnancy because they were able to exercise and relax while isolating with their partner at home. This meant reduced worry about concerns such as the risk of COVID-19 to the health of their baby.

> *“I would say on the pregnancy, I think it had a positive impact on my mental health. I think that I always would have been anxious, I always would have been anxious about the health of the baby, but the fact that I could work from home and that I could exercise every day, and I could eat my own food, in my own house, and that I didn’t have to go anywhere*.*” p12, aged 35-40, 1*^*st*^ *baby*

## 2. Importance of relationships that support coping and adjustment

Having supportive interactions with social contacts had a pronounced impact on mental health for participants in the study, and the absence of support contributed to further feelings of loneliness and isolation during the pandemic.

### 2.1 More time to build a connection as a family

Lockdown restrictions meant that participants were able to spend more time with their partners and children that they may not have previously had the opportunity to do, due to short parental leave allowances.

> *“I mean the only one positive thing for me out of this whole COVID thing… Was that I could spend time, I got to spend a lot of time with my children at this age where they’re talking and really engaging. Because maternity leave you have the baby and then you go back to work when all the exciting stuff starts to happen. So, it was really nice to be at home and spend that time with the children. Because you would never get that opportunity again. But that is the only thing for me that was positive. There’s nothing else at all*.*” P3, aged 35-40, 3*^*rd*^ *baby*

Participants said that this extra time together helped families to re-evaluate their priorities and for some, created a sense of achievement over having bonded during a time of uncertainty and upheaval.

> *“It has made us reassess what’s important and neither me or my husband think that we’re ever going to go back to work in an office full time. He might go maybe three days a week, I might go two. It makes it cheaper for nursery*.*” P6, aged 35-40, 1*^*st*^ *baby*

Social distancing regulations during pregnancy meant having more time to spend at home with a supportive partner, which in turn, benefited their mental health.

> *“The lockdown and the pandemic has meant that my partner’s working from home, and the support that he provides me, I wouldn’t have had if he was at work. So, swings and roundabouts, I guess, a little bit, in terms of not getting support, maybe, from my wider social network, but having the support of my partner has been invaluable*.*” P13, aged 35-40, 1*^*st*^ *baby*

### 2.2 Benefits of the support bubble system

Prior to introduction of the “support bubble” system in the UK, some participants described feeling stressed about increased childcare responsibilities and household chores, leading to stress and frustration about social distancing restrictions.

> *“It just seemed incredibly unfair that my husband was allowed to go to work, we had a new baby, and go and do all of these dangerous thing. But I couldn’t have my mum round for a coffee to help me out*.*” P3, aged 35-40, 3*^*rd*^ *baby*

One participant described her experience after a miscarriage, where she felt further isolated by social distancing restrictions. She had decided to see a friend face-to-face before the bubble system was introduced, out of a perceived need for emotional support.

> *“One of my friends who had actually had a miscarriage herself a few weeks earlier did come over and dropped off some medication and sat with me in the garden and had a chat. Which I don’t know if that was technically allowed then but I wasn’t really thinking about that*.*” P5, aged 30-34, no children, experience of a miscarriage during pandemic*

The introduction of the “support bubble” system in the UK was described as a “lifeline” that made “a huge difference” to the mental health of those who needed the support of close friends and family members.

> *“From December, the rules were that you could form a support bubble if you had a child under 1. And I just think that should have come in so much sooner ‘cause you just need that support – you know, me and my mum have never been really close, but when I became pregnant I suddenly really needed her*.*” P9, aged 35-40, 1*^*st*^ *baby*

### 2.3 Importance of parent groups for support

Most participants described parent groups as an essential source of support to cope with challenges during the pandemic. Although some parent groups had been cancelled, others moved to remote delivery, including breastfeeding classes, National Childhood Trust (NCT) groups, postnatal fitness, and baby singing, sensory, yoga and massage classes.

> *“I was really lucky to attend a lockdown baby group at our children’s centre, so even, obviously, when we had the restrictions, at least that didn’t stop. So, that was good, being able to at least, to me, to just have that one-hour space to bounce off one another, which we did*.*” P11, 35-40, 1*^*st*^ *baby*

Some participants however experienced online groups as unhelpful and found it difficult to build a sense of intimacy with other group members through a computer screen.

> *“I did some of the things online, but most of it, I just thought, you know what, I’d rather be there in person. Socially, not having those groups meant that my mental health was worse than it would have been, I think, otherwise*.*” P13, aged 35-40, 1*^*st*^ *baby*

Many participants, particularly first time mothers, were disappointed that they were unable to make new friendships with other parents due to a lack of face-to-face group options.

> *“We did NCT over Zoom. I can’t say I found it that useful, I found it quite difficult to form the relationships with the other people over Zoom*.*” P9, aged 35-40, 1*^*st*^ *baby*

## 3. Missed pregnancy and parenthood experiences

Although participants reported benefits from some elements of the pandemic, such as being able to work from home with a supportive partner, there were also downsides to social distancing that participants said they found challenging. Many of these issues were more likely to be salient for first-time mothers.

### 3.1 Grief for a missed pregnancy experience

Many participants said they had looked forward to pregnancy rituals like baby showers or shopping for new items for their baby and were disappointed they missed these experiences because of shop closures and stay-at-home orders.

> *“Weird little things that I was really quite looking forward to, like a baby shower*.*…pram shopping, it feels so superficial, but it’s actually quite a nice ritual to go through. It’s like going to pick out baby clothes and find the pram and look at cribs, we never really got to do that*.*” P6, aged 35-40, 1*^*st*^ *baby*

Some participants described sadness about “missing out on” a pregnancy experience that they had wanted or expected because of pandemic-related restrictions.

> *“I’ve waited 37 years to have a baby. I had one in the middle of a pandemic and all the normal things were taken away from me. I felt really sad about it, and scared*.*” P9, aged 35-40, 1*^*st*^ *baby*

### 3.2 Sadness that pregnancy and parenthood could not be shared with others

A number of participants said they felt sadness that important people in their lives missed physically seeing their pregnancy.

> *“I think it was quite hard, actually, being pregnant for the first time and then not seeing people, and people not seeing my bump growing and things like that. I think that was quite a big mental impact on me. And I think it felt like, I don’t know, it wasn’t happening, in a way. It was a bit strange. There were obviously pros to being at home, but I think not seeing people and not having that normal journey through your pregnancy, socially*.*” P15, aged 30-34, 1*^*st*^ *baby*

Not being able to have people visit or to share the experience of being a new parent with others because of the pandemic contributed to feelings of sadness and loneliness.

> *“It kind of felt like a secret, being pregnant, ‘cause I didn’t see anyone, and no one apart from the medical staff and my parents saw me, and it’s kind of like everyone was so caught up in having to adjust with what’s around them and the evolving state of the world, that I kind of felt forgotten about*.*” P8, aged 25-29, 1*^*st*^ *baby*

Some participants said they also felt disappointed that their baby missed seeing other people in their first weeks or months of life because of social distancing restrictions.

> *“I haven’t been able to meet up with my friends and family and share early motherhood with them. Our babies changed so quickly, don’t they, that I feel I’ve had to grieve on what I’ve missed out on and what he’s missed out on*.*” P8, aged 25-29, 1*^*st*^ *baby*

## 4 Mental health consequences of birth partner and visitor restrictions

Being unable to use maternity services with a partner was the most commonly discussed change in service delivery during the pandemic that was experienced as unsettling or stressful. The impact of a partner’s absence was described as most salient, as meaningful experiences were lost and decision-making made more difficult.

### 4.1 Upset about partners being excluded from the healthcare experience

A majority of participants reported that their partners were unable to accompany them to some or all hospital appointments and described this at times as “*stressful”, “difficult” and “traumatising*”. This was especially so for participants in the late stages of pregnancy or experiencing miscarriage. The absence of a partner in times of stress meant that some participants missed moral support and sources of reassurance.

> *“I still had to go into the hospital [after miscarrying] but I would have to go in and go through that process alone and my husband wasn’t allowed into the hospital or into the waiting room even. So, that was really awful because I was not in a great state… The whole thing you’re going through when your body is losing something as well, it’s so traumatic on your body*… *Going to the hospital then alone and being alone through that process and coming out of there alone… It just isolates the two of you even more which is the last thing you need really*.*” P22, aged 35-40, 1*^*st*^ *baby + miscarriage during pandemic*

Participants with high-risk pregnancies, previous miscarriage, or first-time parents were more likely to say that not having their partner present during scans was upsetting.

> *“From my perspective actually, it was just a bit limited in terms of face-to-face time, whereas for him there was nothing, he wasn’t involved essentially, that was probably the worst thing about it. It’s our first baby…” P17, 30-34, 1*^*st*^ *baby*

Participants recalled that missing certain scans was more meaningful at particular stages of development than others. For instance, learning about the sex of the baby, confirming absence of medical conditions (i.e., gestational diabetes) and hearing the heartbeat were described as the hardest appointments to experience alone. Some hospital staff tried to recreate this experience by allowing phone calls, video calls, and sound wave recordings to include partners remotely and as much as restrictions would allow.

> *“In some hospitals they weren’t allowed to tell you what sex the baby was, they couldn’t write it down or anything in case they contaminated the paper, they didn’t take that approach in our hospital. So, the guy that was doing my second scan, the 20-week scan, he did write down the baby’s gender so that we could open it together, so my husband then got to be there for that, so that was lovely*.*” P20, 35-40, 1*^*st*^ *baby*

Not all hospitals however permitted phone calls or recording for women to share the development of their baby with their partner remotely and many participants found this experience upsetting.

> *“I said, ‘can I ring my husband, whilst we’re in the appointment?’ And they were like, ‘you can’t video call’… Okay, that’s weird, but if that’s what you want. They said, ‘can you confirm you’re not going to record us?’… I remember being so insulted. Why would I record you, and even if I did, what skin is it off your nose?” P7, aged 35-40, 2*^*nd*^ *baby*

For some who had to wait alone in hospital for extended periods for active labour to begin, the absence of support and reassurance from their birth partner was disconcerting.

> *“That was really upsetting, being on the maternity ward myself, my waters breaking, getting really scared because they were doing a staff turnover when it all happened and I couldn’t get anyone’s attention… if he’d been there he could have gone off and got someone for me, and it was all just a blur really… And it’s such a traumatic thing for your body to go through isn’t it, giving birth, but, you just need your birth partner there. Whether that’s your partner, or your mum, or your friend, whatever, just someone to bring you down to that level of coping – it’s really important, and that’s what so many women have missed out on*.*” P8, aged 25-29, 1*^*st*^ *baby*

In some cases, parts of the birth were still missed by partners because the labour progressed quickly, or members of staff were unavailable to facilitate birth partner entry to the ward.

> *“I’d been admitted to this observation ward. And then, because they didn’t have a midwife available, whether that is because of COVID or what, I couldn’t be put on the labour ward initially. I did a lot of it on my own, because he couldn’t come up*.*” P21, aged 30-34, 1*^*st*^ *baby*

Some participants who stayed in hospital after the birth of their child reported feeling distressed by not being able to have birth partners or family visit. This was particularly stressful for participants with pre-existing health conditions and post-birth complications.

> *“So, from the Monday onwards, I couldn’t have any visitors, so obviously, that was then challenging … my partner ended up outside the hospital, being told he couldn’t come in, which was very traumatising on both parts*.*” P11, 35-40, 1*^*st*^ *baby*

### 4.2 Stress of decision-making and help seeking without partners present

For those facing pregnancy complications, participants reported that digesting complex medical information and making decisions about the future was inhibited by not being able to have a support person or birth partner present during prenatal appointments.

> *“She had reduced movements once and I had to go into triage by myself. And then, when I was 39 weeks and she wasn’t moving again, I had to go in and they were like, well, we would recommend induction. I had to go and find [my birth partner] outside in the hallway and have a conversation. He couldn’t be in there to talk to the doctor about a really pretty important decision that we had to take. I think he felt a little bit bulldozed by it. Because I’d heard all the information, but then, of course, I couldn’t really relay it*.*” P6, aged 35-40, 1*^*st*^ *baby*

Several participants described feeling unable to advocate for themselves during interactions with hospital staff without the support of a partner.

> *“And the other thing was not having your partner there for any of the antenatal clinics or any of the times I had to go into hospital for check-ups*… *And I think that it was quite hard to make choices on things by myself. And quite hard to have a voice. And I think that your birth partner or your partner throughout the whole thing, it’s quite important to have them there for you to have that voice, because sometimes, when you’re in pain or you’re upset or you’re feeling a bit vulnerable, you can’t actually articulate what you really need or what you want*.*” P15, aged 30-34, 1*^*st*^ *baby*

Consequently, some participants said they paid for private care or changed their birth plan to be able to have their partner’s present with them.

> *“I was really scared I’d be there for days on my own in pain without my husband, or that it would suddenly happen really quickly and he wouldn’t be there, and he’d miss the birth of the baby. So it was quite a major factor in me deciding to ask for a C-section*.*” P9, aged 35-40, 1*^*st*^ *baby*

The presence of birth partners on the postnatal ward was described as particularly important because they provided additional advocacy support and facilitated help seeking at times when participants felt overwhelmed or distressed.

> *“So, the whole, actual giving birth experience, the medical bit, was great, and then the post-natal bit was just awful, it was so horrible and frightening…I realised that I was hallucinating… I couldn’t sleep, because, not only was my baby awake, but all the others were, at various points, as well… on the second day my husband came to visit, I got him to go and say, look, my wife has mental health issues, please can you give her more support*.*” p12, aged 35-40, 1*^*st*^ *baby*

## 5. Maternity services under pressure

Participants recognised the unprecedented circumstances that the pandemic placed on healthcare; however, some expressed frustration with standard elements of care they received that were “exaggerated by the pandemic.”

### 5.1 Emotional impact of delays, staff shortages and service cancellations

Many participants felt the impact of COVID-related service disruptions acutely during pregnancy complications, experiences of miscarriage, and when giving birth.

> *“… You can’t even sit in a waiting room with other people. You’re waiting for a slot to see somebody and they were all so busy in the hospital that you could be in a waiting room for such a long time on your own just waiting to be seen by someone*.*” P22, aged 35-40, 1*^*st*^ *baby + miscarriage during pandemic*

Some described noticing that either staff were absent due to sickness or facing higher workloads than usual leading to some participant saying they felt uncared for.

> *“Lots of staff were off sick, and I think the hospital was in a state of chaos… the hospital were under a lot of stress, but it was just the post-natal stuff…the bit about caring for the baby, there was just no help at all, really*.*” p12, aged 35-40, 1*^*st*^ *baby*

Staff shortages combined with birth partner and visitor restrictions meant that some participants felt a lack of support after childbirth.

> *“I was very disappointed by the level of care that was unfortunately provided to me, because it didn’t help that I couldn’t have anybody there, so there was just, obviously, me and baby, and I felt that the basic needs, like making sure that they’ve given me a bed bath, or support me to go and use the shower or supporting me to get changed, or any of that, just didn’t happen, whatsoever*.*” P11, 35-40, 1*^*st*^ *baby*

Consequently, some worried that the antenatal care they were receiving was not of a standard they had expected, or that steps to protect their health or their baby’s health were being missed.

> *“I felt like, to some extent, they might not be following up things they would normally follow up or perhaps dealing with things with the same urgency that they normally would, because of COVID*.*” P15, aged 30-34, 1*^*st*^ *baby*

At a time where many felt lonely, unsupported and vulnerable, some participants said they felt guilty for asking staff for help or information.

> *“I remember contacting the assisted conception unit in the hospital and them saying, ‘it’s really difficult here, we’re short-staffed and we’ve got COVID happening, this is the emergency*.*’ So, you feel really bad about asking for support. You realise that no one’s dying by at the same time, oh my God, emotionally you feel like you’re dying. I know that’s quite dramatic to say but you feel so low and then you feel like you can’t contact anybody. So, it compounds that feeling*.*” P22, aged 35-40, 1*^*st*^ *baby + miscarriage during pandemic*

### 5.2 Staff seemed at times unaware of social distancing rules

Several participants described interactions with hospital staff where COVID-related rules were unclear or implemented inconsistently at various points along the care pathway.

> *“I went by myself, and the entrance, it said, if this is your first appointment at the foetal medicine unit, your husband can come, your partner can come. I asked them, can my partner come? They were, we don’t know, they keep changing the rules, none of us has got any idea. So, they went and asked, and in the end they were like, just send him up*.*” P7, aged 35-40, 2*^*nd*^ *baby*

Several aspects of maternity service care, including lack of clarity about the hospital social distancing rules and absence of partners for support, meant many participants made additional effort to gather information and advocate for their wants and needs during appointments.

> *“I think there was some confusion with the doctors and the nurses around what the policies were, not that they told me that, but that’s what I felt. And then, when I had more of an understanding of what the policies were, when I was a bit more, like, this is what I’m allowed to do, then I think they gave in a bit more*.*”* P13, aged 35-40, 1^st^ baby

## 6. Lack of connection with staff

The subject of connection was discussed in relation to trust, touch and support from staff, almost interchangeably by some of the participants in the study. These factors were described as important in terms of satisfaction with care received.

### 6.1 Communication difficulties

Participants reported some communication difficulties with staff during their interactions with maternity services. Several participants said they found it difficult to communicate with healthcare staff whilst wearing protective equipment during appointments.

> *“That was really weird, and just going in with masks and seeing the doctors and the nurses through masks, that was all really weird. I’m quite a social person and I chat, it just made it all a lot more difficult*.*” P13, aged 35-40, 1*^*st*^ *baby*

Some participants felt that they did not receive the same quality of maternity service care during the pandemic compared to pre-pandemic times. This was a concern as some participants felt elements of their care might be missed due to remote consultations, particularly for those having their first child and uncertain of what to ask for or what was considered “*normal*.*”*

> *“And I know that, normally, the health visitors would see you once a month… that is something that I really felt was quite a worry for me, especially in the beginning, because my son was premature and I was concerned about his weight and concerned about just lots of things developmentally. And I think just having a phone call about that was quite concerning*.*” P15, aged 30-34, 1*^*st*^ *baby*

Many found it difficult to “*build a relationship*” and gain “*reassurance*” from midwives and consultants over the phone. Participants with pre-existing mental health concerns, pregnancy-related anxiety, experiences of miscarriage and pregnancy complications were more likely to say the lack of face-to-face care was a source of concern.

> *“The pregnancy didn’t show very visibly on me… I had no bump at all, really, for about five and a half months. So, I was always quite anxious, ‘is the baby developing properly?’… because all the midwife appointments were on the phone, it was probably only about five months in, where I actually got measured, and they were like, oh, yes, that’s fine*.*” p12, aged 35-40, 1*^*st*^ *baby*

Several participants said that health issues for themselves and their baby were missed because of a lack of face-to-face appointments and physical examinations during COVID-restrictions.

> *“Post-natal, I think it’s a six-week check for mum. That didn’t happen in person, and that was, for me, a really big issue, because my C-section scar was infected… nobody was able to check it after, to make sure that it was okay, and it would have normally happened at the six-week appointment. But because that happened over the phone, they weren’t able to have a look at it. So, I think that was, personally, that should have been an appointment that happened face-to-face*.*”* P13, aged 35-40, 1^st^ baby

### 6.2 Prevention of touch due to COVID-related restrictions

Due to COVID-restrictions, participants reported that many healthcare staff were unable to provide hands on care. Experiences of staff being unable to touch participants and their babies had an impact on how participants felt about their care on the postnatal ward and beyond.

> *“I think it was just difficult in terms of, some of the midwives in the hospital, the advice is not to touch the babies so much. And I think when we were at home, you’ve still got someone coming in, and wearing a mask. It couldn’t be as personal, maybe, or interactive as what it might have been*.*” P22, aged 35-40, 1*^*st*^ *baby + miscarriage during pandemic*

The rules around lack of touch were particularly upsetting for participants staying on the postnatal wards. When staff were unable to provide one-to-one physical care coupled with birth partner visiting restrictions, many participants felt unsupported, stressed and alone.

> *“[My baby] was crying, and none of the nurses were able to pick him up. I was pulling on my trousers, and I had him in my hand, and I couldn’t even pass him over to a doctor, to anyone, so I had to put him back in the buggy, and he was crying, I was trying to change. It was just complete madness, and you could see that the nurses were looking at me, quite sympathetic, but they couldn’t do anything*.*” P13, aged 35-40, 1*^*st*^ *baby*

### 6.3 Disruptions to continuity of care

Pandemic-related pressures on the health service affected continuity of care and participants said that this compounded their feelings of being alone, particularly for first time parents and participants with a high-risk pregnancy.

> *“I had to tell my story every time, that was just really distressing, and none of them read the notes in advance…I cannot describe how stressful my pregnancy was, and it was definitely compounded by having no-one hold my hand through it. And of course, now, they’ve got a policy of the same midwife for the whole pregnancy. And I’ve seen that come a bit out of the pandemic, and a bit out of people’s feedback in general*.*” P7, aged 35-40, 2*^*nd*^ *baby*

The impact of frequent staff changes and lack of staff availability meant that some participants felt less supported than if they had seen the same health professional throughout their pregnancy.

> *“I rarely saw my actual, my allocated midwife. Each time I went, it was someone different… I couldn’t build that rapport when it’s not the same person every time. So, yeah that was tough*.*” P8, aged 25-29, 1*^*st*^ *baby*

As the pandemic progressed and some services adjusted their practices, being able to see the same member of staff helped to build trust and increase feelings of reassurance.

> *“I would say from 36 weeks onwards, I was seeing a midwife nearly every week, and I was seeing the same midwife, and that really made a difference, with a student, who was really good as well. I felt far more supported, because I was like, I’m seeing, I know the midwife’s name, I’m going to her next week, I’ll save up this question. She’s making sure that everything’s okay, she’s feeling the baby move, listening to his heartbeat, all that kind of stuff, and I felt far more assured*.*” P4, 30-34 1*^*st*^ *baby*

## Discussion

In this study, we explored how social distancing restrictions affected the mental health of women experiencing a pregnancy and accessing UK maternity services during the COVID-19 pandemic. Aligned with existing research, the women in our study shared concerns about reduced social contact and support, (3,34) and feelings of loneliness and isolation (35,36) which were exacerbated by the pandemic and associated restrictions. Our findings build on existing research by eliciting the perspectives of women throughout different stages of pregnancy and early parenthood and add further insight into how loneliness and isolation was felt by pregnant women throughout the COVID-19 pandemic. Isolation was experienced by participants at multiple points along the antenatal care pathway, including having continual remote consultations with pre- and postnatal care staff, being unable to see the same members of staff during their pregnancy, and being unable to have birth partners present during their interactions with services.

At the start of the pandemic, experts recommended precautions to reduce the risk of COVID exposure during antenatal visits, including offering women with uncomplicated pregnancy remote appointments.(14) While remote healthcare appointments may have increased access to healthcare among some groups during the pandemic, offering appointments remotely in the future is not a preference experienced by all, particularly for those with serious health concerns.(37) Qualitative research interviews conducted in Canada suggest that women may prefer virtual postnatal care because it helps to regulate their family routine, reduce stress and save on expenses associated with travel to consultations. (38) However, for women in our study experiencing a first pregnancy, pregnancy complications or pre-existing health conditions, remote care was a source of concern, leading to feelings of uncertainty and increased stress.

The perceived lack of connection reported between people experiencing pregnancy and their midwives is important because it can prevent trust and empowerment, (39) as described by participants in this study. Previous studies have shown that women are more likely to feel as though their needs are being met and their voices heard when a relationship exists between themselves and midwifery staff.(39) For women in our study with high-risk pregnancies who may have been more vulnerable to isolation, grief and fear (40), relationships with health professionals were reported to be especially important. Lack of continuity of care and trust has been reported by women prior to the COVID-19 pandemic when using UK maternity services (41), but participants in this study said that this was exacerbated by COVID-related restrictions, such as mandatory use of PPE, remote appointments, limitations to hands-on physical care, and staff shortages in their health service. Some also said that this disrupted connection with staff alongside partner and visitor restrictions left them feeling alone during childbirth and postnatal care.

Participants in our study described the various ways in which their social ties were cut during the pandemic due to social distancing restrictions but also pandemic-related cancellations in parent groups and community services. These restrictions and cancellations compounded feelings of loneliness and isolation and ultimately mental health and wellbeing. Taken together with existing research,(42,43) these findings highlight the importance of peer support groups for parents in times of pandemics. Introduction of the support bubble system whereby one household could form a support network with one other household (44) was a key change in social policy that brought about noticeable differences for participants struggling with isolation, miscarriage, new motherhood, childcare demands and adverse mental health. For other groups, including domestic abuse survivors (45) and parents with young children (32) many have also gained important sources of social support resulting from this policy. However, bringing these support bubbles to women earlier during pregnancy (rather than just post-birth) could have helped to mitigate further issues reported in this study.

Social distancing regulations have been reported as psychologically challenging among different groups within the general population (46,47), and participants in our study reported aspects of their social distancing experience during the pandemic that threatened their mental health and wellbeing. However, there were pandemic-related changes that were also helpful and meaningful. For those in the early stages of pregnancy, being required to stay at home helped them to feel safe from the virus and better able to manage their pregnancy symptoms. It is notable that some participants reported having to take less time off work due to being able to work and manage their symptoms better from home, suggesting that more flexible policies on working from home during pregnancy could reduce sick days amongst pregnant women. Having partners at home at the same time also supported feelings of wellbeing by increasing access to practical and emotional support at home. A consistently reported finding is the strengthening of families (3) and relationships (36) under lockdown restrictions, but we found this connection also helped families make decisions about their future and feel more stable in times of great uncertainty during the pandemic.

### Strengths and limitations

Owing to in-depth qualitative interviewing methods and a data collection period that spanned three national lockdowns in the UK, we were able to present a wide range of detailed experiences throughout the various stages of the COVID-19 pandemic. Remote interview methods meant that women across the UK could take part in the study; however, people without access to the internet may have been inadvertently excluded from taking part. Although we attempted to sample a wide range of demographic characteristics to explore the impact of these factors on women’s experiences of pregnancy during the pandemic, our sample was restricted to married women, who are highly educated, in fulltime employment and aged mostly over 30, with some suspected but no confirmed COVID-19 diagnosis. Consequently, we were unable to “compare and contrast” the cases presented here as we might have been able to, had the sample been more demographically diverse.(48) Characteristics such as gender identity (49), marital status, age, ethnicity and socioeconomic group (50) can compound experiences of isolation and marginalisation in pregnancy, and therefore, we recommend future researchers explore the impact of pandemic-related changes among these specific communities.

## Conclusions

Our findings highlight aspects of care that must be taken into consideration in pandemics or disaster-related situations, in order to protect the mental health of people experiencing pregnancy and miscarriage. Specifically, availability of a birth partner or support person must be permitted where possible, as these restrictions brought about the most stress and uncertainty for women in our study. Further, support bubbles not just post-birth but during pregnancy should be explored as a priority to provide adequate support with mental health, physical symptoms, and high-risk pregnancies. Pregnant women in this study said they experienced a loss of social support and access to parent groups during the COVID-19 pandemic, which had a detrimental impact on their mental health and wellbeing, so more development of online or socially distanced support groups could help to address this issue in the future. The pandemic also placed additional pressures on the delivery of maternity services and many women reported dissatisfaction with aspects of care that were exacerbated by social distancing restrictions. Further work is needed to explore the experiences of maternity staff during the COVID-19 pandemic, to identify what further support they feel is needed for maternity services in the future, as we move beyond the pandemic.

## Data Availability

Data from the qualitative research interviews is not publicly available, as this might compromise the identity and anonymity of our research participants.

